# Incidence of perioperative anaphylaxis in 2021 in Japan: Survey of 34 hospitals of Social Welfare Organization Saiseikai Imperial Gift Foundation, Inc

**DOI:** 10.1101/2022.11.08.22282102

**Authors:** Yasuyuki Suzuki, Shuang Liu, Natsumi Yamashita, Naohito Yamaguchi, Yasushi Takasaki, Toshihiro Yorozuya, Masaki Mogi

## Abstract

Perioperative anaphylaxis (POA) is a severe complication with a low incidence and diverse risk factors. Its low incidence makes a detailed survey of POA challenging. Recent large-scale surveys in Japan have focused on identifying the causative agents using the basophil activation test. Only facilities, such as university hospitals, that can dispatch samples for this test participate, which may introduce bias. We surveyed the incidence of POA primarily in secondary hospitals, such as the Saiseikai hospitals, throughout Japan.

We targeted data collection for the year 2021. Thirty-five facilities answered (Supplementary 1, 2); we excluded 1 reporting no surgeries. In total, 70,523 surgeries were performed, with POA diagnosed in 7 cases. For diagnosis, the skin test was used in 3 cases and quantification of histamine and tryptase in 1 case. In 3 cases, diagnosis was based on the time to onset of POA after drug use. Multiple tests are important to ensure patient safety. However, in this survey, no hospital performed in vitro tests to identify the cause. This is major limitation, however, as many hospitals do not have experimental laboratories, we believe that this study, which focuses on secondary care institutions, contributes to the literature significantly by presenting the current situation in clinical practice.

## Introduction

Perioperative anaphylaxis (POA) is a severe complication with a low incidence and diverse risk factors.^1–3^ Medical conditions, lifestyle factors, drug use, and the country of residence affect POA risk. However, these differences vary with time, mandating routine updates to such data. Its low incidence makes a detailed survey of POA challenging. Recent large-scale surveys in Japan have focused on identifying the causative agents using the basophil activation test (BAT). Only facilities, such as university hospitals, that can dispatch samples for this test participate, which may introduce bias.^4, 5^ To our knowledge, no study has included secondary health care providers that do not have an experimental laboratory setup. The Social Welfare Organization Saiseikai Imperial Gift Foundation, Inc. is a group of institutions established for the poor in 1911 by Emperor Meiji. We surveyed the incidence of POA primarily in secondary hospitals, such as the Saiseikai hospitals, throughout Japan. The Ethics Committee approved this study within the Saiseikai Research Institute of Health Care and Welfare.

## Methods

We issued the survey to 56 Saiseikai hospitals. We targeted data collection for the year 2021, gathering data on surgery volume, POA incidence, the causative agent, and diagnostic methods. The diagnostic criteria proposed by the Japanese Society of Allergology were used. Considering surgery volume and POA incidence, a confidence interval with a Poisson distribution was constructed to evaluate incidence per institution.

## Results

Thirty-five facilities answered (Supplementary 1, 2); we excluded 1 reporting no surgeries. In total, 70,523 surgeries were performed, with POA diagnosed in 7 cases.

Regional distribution showed that Kanto, Kansai and Northern Kyushu regions, which have a high population density, witnessed many cases (Fig. 1A). The incidence rates were similar between high-volume and non-high-volume centres (Fig. 1B) although a higher surgery volume should expectedly be associated with a higher POA incidence.

**Fig. 1A.**
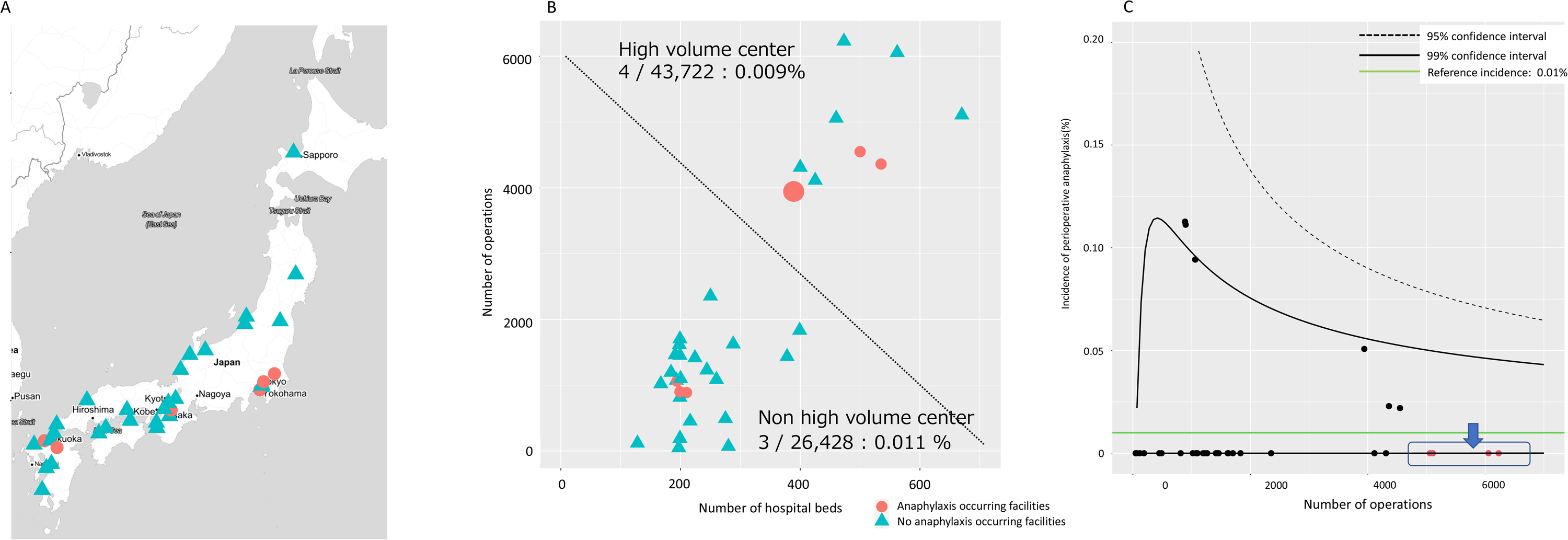
Location distribution of facilities where perioperative anaphylaxis occurred. The healthcare facilities surveyed are distributed not only in urban areas but also in rural areas. Facilities experiencing perioperative anaphylaxis are concentrated in Kanto (e.g., Tokyo), Kansai (e.g., Osaka) and Kitakyushu (e.g., Hakata) regions. These regions have high population densities and hence witness more surgeries, and consequently, more cases of perioperative anaphylaxis. The map was created using the *ggmap* package version 3.0.0 on R version 4.1.3. Red circles: facilities where anaphylaxis occurred. Blue triangle: facilities that did not report anaphylaxis occurrence. **Fig. 1B. The association between the occurrence of perioperative anaphylaxis and the number of beds and surgeries per facility**. Scatter plots visualising whether anaphylaxis is more common in high-volume centres with a high number of operations than in non-high-volume centres. The incidence of both groups was examined using a Fisher exact test, which showed no significant difference (p=1.00). Scatterplot analysis and the Fisher exact test were performed using R version 4.1.3. Red circles: facilities where anaphylaxis occurred. Blue triangle: facilities that did not report anaphylaxis occurrence. **Fig. 1C. Funnel plot analysis to determine the association between the number of surgeries and the incidence of anaphylaxis**. To determine the effect of the number of surgeries at each centre on the incidence of anaphylaxis, a funnel plot was constructed with confidence intervals based on a Poisson distribution and the mean incidence rate obtained in this study as the reference value. The graph was drawn using R version 4.1.3 with modification in the source code of the program (https://fukui-ke-0507.shinyapps.io/funnel/) published by Dr. Fukui, Hiroshima University (permission obtained). Black plots: within 95% confidence interval. Red plots: outside 95% confidence interval.

A Poisson regression model was applied to the number of POA cases relative to the surgery volume, and a funnel plot was drawn using the reference anaphylaxis incidence rate of 0.01% for in this study. Thirty facilities fell within the 95% confidence interval (CI). The facilities falling outside the CI reported no POA events despite conducting a high volume of surgeries, although clinically insignificant (Fig. 1C).

The most suspected causes were the use of rocuronium (3 cases), sugammadex (2 cases), latex (1 case), and an angiography contrast agent (1 case). For diagnosis, the skin test was used in 3 cases and quantification of histamine and tryptase in 1 case. In 3 cases, diagnosis was based on the time to onset of POA after drug use.

## Discussions

A study that included tertiary hospitals in Japan reported a POA incidence of 0.036%.^5^ National Audit Projects 6 (NAP6) also reported an incidence of about 1 in 10,000 cases.^3^ The incidence across the Saiseikai hospitals is in line with that in previous reports. Funnel plot analysis showed no effect of surgery volume on incidence. Therefore, we hypothesise that POA incidence is not significantly affected by differences in the country of residence, the region, and hospital size. However, incidence can change over time and because of trend changes in drug use.

The most common cause was rocuronium use, as corroborated by a previous report.^4^ POA because of rocuronium use manifested not only as a type I allergy with specific IgE antibodies, but also through histamine release owing to the stimulation of Mas-related G protein-coupled receptor X2 in mast cells.^6^ Hence, the use of this high-risk agent should be carefully considered.

Sugammadex is an excellent drug that specifically adsorbs rocuronium and is considered a high-risk agent in Japan owing to its frequent use.^4, 5^ Sugammadex’s cyclodextrin structure is widely used in daily life products, such as deodorants and diet food, which implies a high sensitisation potential.

According to the guidelines, prompt diagnosis of POA based on physical observations is critical to its management in the acute phase.^7^ Equally important are objective tests, such as blood tests, for definitive diagnosis. In type I allergy, histamine and tryptase are quantified for diagnosis. However, in Japan, only few outsourcing laboratories quantify these biomolecules for research purposes.

Identifying the causative agent is vital for patient safety. Although skin testing is the most basic approach and is considered safe, it is a challenging test and hypersensitivity reactions occur in some cases.

Specific IgE antibody detection is a safe in vitro testing method that could diagnose type I allergy; however, it is expensive, particularly in case of uncommonly used drugs. In the BAT, a suspect drug is added to basophils collected from patient blood, and changes in cell surface markers are observed. BAT requires active basophils within 4 hours of blood collection, limiting its use to facilities with flow cytometers.^8^

The mast cell activation test (MAT) uses cultured mast cells bound to serum IgE antibodies and, similar to BAT, detects cell surface marker changes. However, maintaining adequate activity in cultured mast cells requires further study. Furthermore, the response may vary with the cell line.^9^

Considering the possibility of false-negatives, and as NAP6 indicates, multiple tests are important to ensure patient safety.^3, 10^ However, in this survey, no hospital performed in vitro tests to identify the cause, because these tests are difficult for local hospitals.

A limitation of this survey is its small sample size of 7 cases despite it being a multi-centre survey. Furthermore, this study had a retrospective design and is not as extensive as the previously mentioned prospective studies. However, as many hospitals do not have experimental laboratories, we believe that this study, which focuses on secondary care institutions, contributes to the literature significantly by presenting the current situation in clinical practice.

In future studies, we plan on devising an examination that can facilitate identifying the causative agent in secondary healthcare facilities. In particular, using MAT with patient serum that can be stored for a long time and low-cost quantification of histamine and tryptase, will be studied.

## Data Availability

All data produced in the present study are available upon reasonable request to the authors.

## Acknowledgments

We thank Mr. Yuji Mochida, a data analysis professional at the Saiseikai Research Institute. We also thank Ms. Yuko Yoshizumi, who supports our research at the same institute.

## Author contributions

Writing the paper: Y.S.

Data analysis: Y.S.

Planning analysis: Y.S., N. Yamaguchi.

Advice the date analysis: N. Yamashita., Y.T.

Supervision: L.S., T.Y., M.M.

## Declaration of interest

The authors declare that they have no conflicts of interest.

## Funding

Y.S. was supported by the Japan Society for the Promotion of Science (JSPS), Grants-in-Aid for Scientific Research KAKENHI grant number 21K16549.

## Figure legends

**Supplementary 1**

List of healthcare organisations that cooperated with our survey. Special thanks to all staff who responded.

**Supplementary 2**

A list of hospitals grouped by region. The number of beds and the number of surgeries per year, the number of anaphylactic incidents, the suspected causative drug and the test method used for diagnosis are shown. Hospital numbers are given in the corresponding order in which they were reported.

## Notes

### Competing Interest Statement

The authors have declared no competing interest.

### Author Declarations

The Ethics Committee approved this study within the Saiseikai Research Institute of Health Care and Welfare.

