## Supplementary 1 for "Incidence of perioperative anaphylaxis in 2021 in Japan: Survey of 34 hospitals of Social Welfare Organization Saiseikai Imperial Gift Foundation, Inc"

We wish to acknowledge the staff of the following medical centres for their cooperation in the survey.

Saiseikai Karatsu Hospital, Saga, Japan

Isikawaken Saiseikai Kanazawa Hospital, Ishikawa, Japan

Saiseikai Toyama Hospital, Toyama, Japan

Higasikanagawa Rehabilitation Hospital, Kanagawa, Japan

Saiseikai Fukuoka General Hospital, Fukuoka, Japan

Saiseikai Misumi Hospital, Kumamoto, Japan

Saiseikai Kanagawa Hospital, Kanagawa, Japan

Saiseikai Niigata Hospital, Niigata, Japan

Saiseikai Yokohamashi Nanbu Hospital, Kanagawa, Japan

Saiseikai Gose Hospital, Nara, Japan

Okayama Saiseikai general Hospital, Okayama, Japan

Kitakami Saiseikai Hospital, Iwate, Japan,

Saiseikai Nara Hospital, Nara, Japan

Kagawaken Saiseikai Hospital, Kagawa, Japan

Saiseikai Nakatsu Hospital, Osaka, Japan

Fukuiken Saiseikai Hospital, Fukui, Japan

Ryugasaki Saiseikai Hospital, Tochigi, Japan

Saiseikai Kumamoto Hospital, Kumamoto, Japan

Saiseikai Wakayama Hospital, Wakayama, Japan

Saiseikai Otaru Hospital, Hokkaido, Japan

Saiseikai Arita Hospital, Wakayama, Japan

Niigataken Saiseikai Sanjo Hospital, Niigata, Japan

Yamagutiken Saiseikai toyoura Hospital, Toyama, Japan

Saiseikai Fukusima General Hospital, Fukushima, Japan

Saiseikai Iizuka Kaho Hospital, Fukuoka, Japan

Saiseikai Yokohamasi Toubu Hospital, Kanagawa, Japan

Saiseikai Imabari Hospital, Ehime, Japan

Saiseikai Yahata General Hospital, Fukuoka, Japan

Oita Prefecture Saiseikai Hita Hospita, Oita, Japan

Saiseikai Sendai Hospital, Miyagi, Japan

Shimaneken Saiseikai Gotu General Hospital, Shimane, Japan

Tokyo Saiseikai Central Hospital, Tokyo, Japan

Saiseikai Kyoto Hospital, Kyoto, Japan

Saiseikai Moriyama Municipal Hospital, Shiga, Japan

Saiseikai Matuyama Hospital, Ehime, Japan
