## Supplementary 2 for "Incidence of perioperative anaphylaxis in 2021 in Japan: Survey of 34 hospitals of Social Welfare Organization Saiseikai Imperial Gift Foundation, Inc"

| Hospital | Number of operations | Number of beds | Number of anaphylaxis | Methods of diagnosis | Causative substance |
| --- | --- | --- | --- | --- | --- |
| Kyushu | | | | | |
| S\_01 | 1074 | 193 | 0 | NA | NA |
| S\_04 | 3941 | 390 | 2 | Based on timing of drug use and physiological findings | Latex, Angiographic contrast agents |
| S\_05 | 118 | 128 | 0 | NA | NA |
| S\_17 | 4311 | 400 | 0 | NA | NA |
| S\_24 | 47 | 197 | 0 | NA | NA |
| S\_27 | 1833 | 399 | 0 | NA | NA |
| S\_28 | 899 | 199 | 1 | Skin test | Rocuronium |
| S\_29 | 1229 | 244 | 0 | NA | NA |
| Hokuriku | | | | | |
| S\_02 | 1085 | 260 | 0 | NA | NA |
| S\_03 | 2354 | 250 | 0 | NA | NA |
| S\_07 | 4114 | 425 | 0 | NA | NA |
| S\_15 | 5058 | 460 | 0 | NA | NA |
| S\_21 | 814 | 199 | 0 | NA | NA |
| Kanto | | | | | |
| S\_06 | 1703 | 199 | 0 | NA | NA |
| S\_08 | 4549 | 500 | 1 | Quantification of histamine and tryptase. | Rocuronium |
| S\_16 | 887 | 210 | 1 | Based on timing of drug use and physiological findings | Rocuronium |
| S\_25 | 6056 | 562 | 0 | NA | NA |
| S\_31 | 4361 | 535 | 1 | Skin test | Sugammadex |
| Kansai | | | | | |
| S\_09 | 1020 | 167 | 0 | NA | NA |
| S\_12 | 1061 | 194 | 1 | Skin test | Sugammadex |
| S\_14 | 5107 | 670 | 0 | NA | NA |
| S\_18 | 1100 | 200 | 0 | NA | NA |
| S\_20 | 1195 | 184 | 0 | NA | NA |
| S\_32 | 1625 | 288 | 0 | NA | NA |
| S\_33 | 190 | 199 | 0 | NA | NA |
| Chugoku | | | | | |
| S\_10 | 6229 | 473 | 0 | NA | NA |
| S\_22 | 489 | 275 | 0 | NA | NA |
| S\_30 | 68 | 280 | 0 | NA | NA |
| Tohoku | | | | | |
| S\_11 | 1415 | 224 | 0 | NA | NA |
| S\_23 | 450 | 216 | 0 | NA | NA |
| Shikoku | | | | | |
| S\_13 | 1616 | 198 | 0 | NA | NA |
| S\_26 | 1461 | 191 | 0 | NA | NA |
| S\_34 | 1265 | 199 | 0 | NA | NA |
| Hokkaido | | | | | |
| S\_19 | 1433 | 378 | 0 | NA | NA |
